# SGLT2 inhibitor use in type 2 diabetes in England: a population-based cross-sectional study of uptake of NICE guidance

**DOI:** 10.64898/2026.02.04.26343917

**Authors:** Patrick Muller, Jonathan Wray, Muksitur Rahman, James Hawkins, Chirag Bakhai, Daniel J Cuthbertson, Robert Willans, Eleanor Yelland, Shaun Rowark, Magdalena Watras, Luke Sheridan Rains, Amanda I Adler, Lesley Owen

## Abstract

**Aims:** An update to the NICE Type 2 diabetes (T2DM) guideline in February 2022 recommended an SGLT2 inhibitor be offered to people with cardiovascular disease (CVD) or heart failure (HF) as comorbidities and considered for people at high CVD risk. We report uptake of this guideline in England 18 months after its publication.

**Materials and Methods:** Observational cohort study using Clinical Practice Research Data Link records linked to hospital admissions. Presence of a current prescription for an SGLT2 inhibitor was evaluated in people with T2DM on 1^st^ September 2023, stratified by CVD category (CVD only; HF only; both; high CVD risk; low CVD risk) and chronic kidney disease status, and by age, gender, ethnicity, deprivation, and T2DM duration. Adjusted associations between patient characteristics and uptake were evaluated using logistic regression.

**Results:** In the cohort of 587,826 people with T2DM, the percentage with a current prescription was 19.5% for people with CVD, 29.4% for people with HF, 30.5% for people with both CVD and HF, and 19.9% and 20.2% respectively for people at high and low CVD risk. In age-stratified analyses uptake was higher in people with more comorbidities. In adjusted models, uptake was lower in people aged >60, women, Black people, and people living in areas of higher deprivation.

**Conclusions:** Whilst prescribing of SGLT2 inhibitors continues to rise in England, recent trends indicate an opportunity remains to increase uptake. Action is necessary to address inequalities by ethnicity and deprivation, and lower uptake for people with T2DM and CVD without HF.

## INTRODUCTION

The prevalence of type 2 diabetes (T2DM) has increased in England in recent decades, affecting approximately 7% of adults by March 2024.^1^. The percentage of people with T2DM who are living with hypertension, renal disease, and cardiovascular disease as comorbidities has also risen in the same period.^2^ Goals for treating T2DM have evolved from focusing on managing glycaemia to including primary and secondary prevention of cardiovascular and renal disease. Recent trial evidence on the cardiorenal benefits of SGLT2 inhibitors in people with T2DM has led to their addition to treatment strategies to achieve these goals.^3–5^

Between 2014 and 2023, the National Institute for Health and Care Excellence (NICE) in the UK published a range of guidance on SGLT2 inhibitors (Figure 1). In 2022, following Technology Appraisals (TAs) recommending SGLT2 inhibitor therapies for escalation of glucose-lowering treatment in T2DM, for symptomatic heart failure, and for chronic kidney disease (CKD), NICE updated its T2DM guideline.^6^ The update expanded the T2DM population to be offered SGLT2 inhibitor therapy, recommending that an SGLT2 inhibitor with proven cardiovascular benefit be offered for people with heart failure or CVD as a comorbidity, and considered for people at high risk of CVD. The change was informed by network meta analyses indicating 10-40% lower odds of cardiovascular mortality associated with taking metformin and an SGLT2 inhibitor compared to metformin alone.^7^

**Figure 1.**
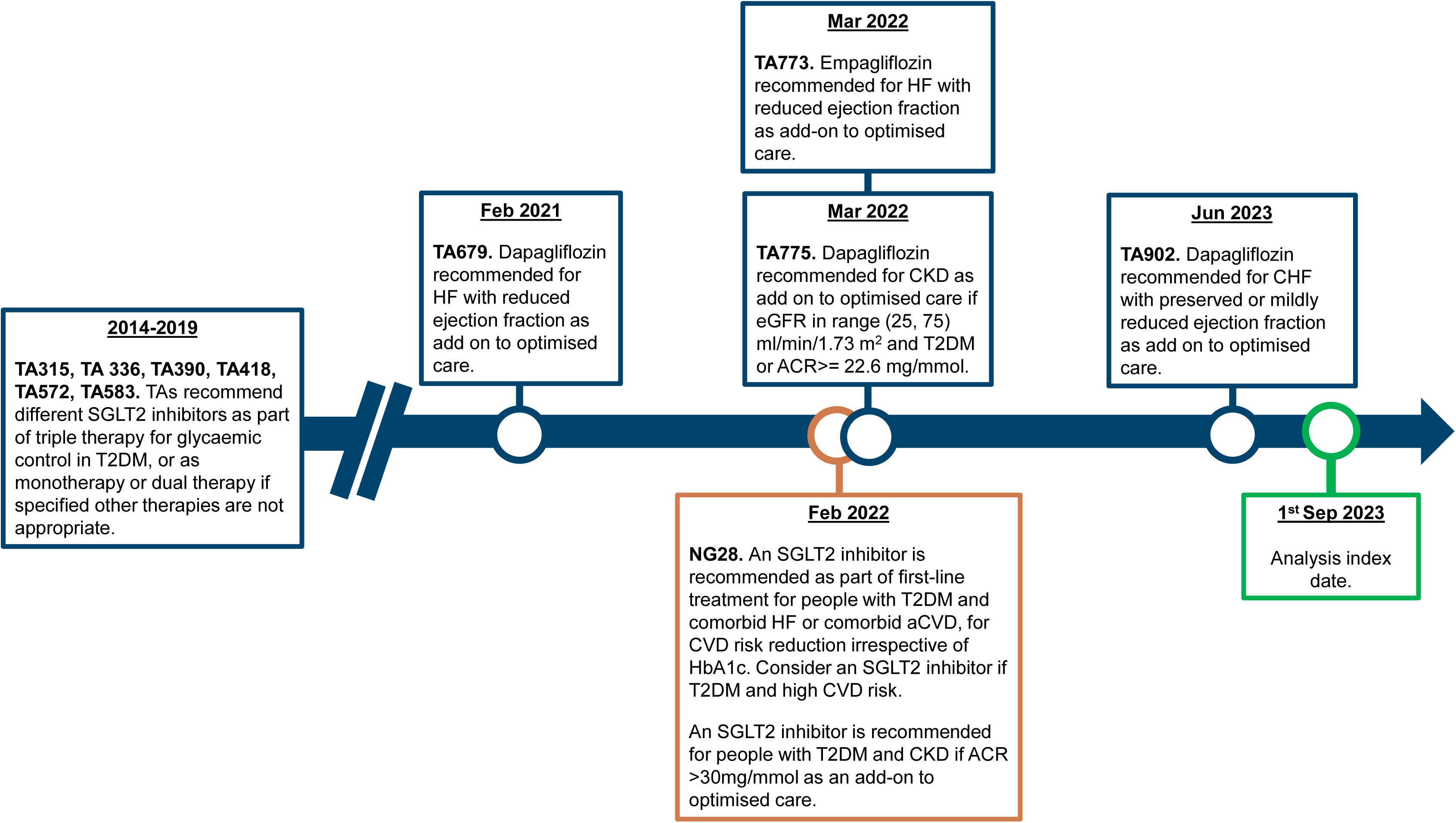
Timeline of selected NICE Technology Appraisals (blue) and guideline updates (orange) recommending SGLT2 inhibitors for different indications, 2014-2023.

NICE prioritises evaluation of the uptake of guidelines and supporting reductions in health inequalities.^8^ The increasing availability of routinely collected electronic health and prescription records is facilitating evaluations of uptake and inequalities in uptake in the populations in scope of NICE guidance.^9^ In the present study, we report variation in the percentage of people with T2DM who had a current SGLT2 inhibitor prescription (whether as new initiators or through treatment switching) on 1^st^ September 2023 according to patients’ comorbidity status, in a large and representative sample of English GP practices. We assess the evidence for inequalities in uptake according to age, gender, deprivation, and ethnicity, separately for patients stratified according to their CVD group (with CVD; with HF; with CVD and HF; high CVD risk score; low CVD risk score) and by whether they have CKD. We consider the clinical and organisational factors that may explain the patterns we report. Finally, we compare our findings to those of other studies, and discuss the implications of our results with reference to the most recent update to the NICE T2DM guideline in 2026.

## MATERIALS AND METHODS

### Data sources

The cohort for analysis was extracted from the Clinical Practice Research Datalink (CPRD) Aurum.^10^ CPRD Aurum includes recorded symptoms, diagnoses, test results, and medication data for people at participating GP practices from 1995 to present, with approximately 24% coverage of the UK population by December 2023.^11^ Patients in CPRD are broadly representative of the wider population with respect to their geographic distribution, deprivation, age, gender, and ethnicity.^12,13^

Patients’ CPRD records were linked to their Hospital Episode Statistics (HES) records, to retrieve diagnoses and procedures recorded in NHS hospital inpatient admissions (with linkage up to March 2021 only),^14^ and to their Index of Multiple Deprivation (IMD) quintile, categorised from 5 (least deprived) to 1 (most deprived). The IMD ranks small areas in England from most to least deprived using a single score based on 7 components of overall deprivation.^15^

Lists of codes from the SNOMED (for CPRD) and ICD-10 and OPCS-4 (for HES) libraries were collated to select demographics, comorbidities, risk factors, and current medicines from the CPRD-HES dataset. Where relevant code lists were available from the General Practice Domain Set maintained by NHS England,^16^ the Open Code Lists website maintained by the Bennett Institute,^17^ or the CVDPrevent audit,^18^ these were used. Otherwise, code lists were taken from the literature or created using the World Health Organisation ICD-10 codes library,^19^ or the CPRD code browser. The codelists are available online (Appendix 1).

### Cohort selection, stratification, exposure and outcome definitions

Patients were selected for the cross-sectional analysis of treatment uptake on 1 September 2023 (the “index date”) through identifying patients with any record of T2DM and no other form of diabetes in CPRD between 1^st^ January 2000 and the index date, and retaining those whose records could be linked to both HES and IMD datasets, who were judged by CPRD to have a ‘research-quality’ record, and who were aged 18 or over on the index date.^20^ People were excluded if they died or de-registered prior to the index date, or if they had less than one year registration on the index date.

The selected people were categorised into mutually exclusive groups of CVD without HF, HF without CVD, CVD with HF, high CVD risk score (QRISK2 score >10%^21^) without CVD or HF and low CVD risk score without CVD or HF. CVD was selected if they had a record of stroke, myocardial infarction, peripheral arterial disease, angina, or atherosclerosis These were further subcategorised according to CKD status, with CKD defined by either an ACR > 3 mg/mmol or eGFR <60 ml/min/1.73m^2^ in the most recent record of these in General Practice within two years prior to the index date. Patients with eGFR > 60 ml/min/1.73m^2^ and without a recent ACR record were categorised as not having CKD. Most people did not have a recent QRISK2 recorded in CPRD Aurum. This was instead estimated using an algorithm developed by researchers at the London School of Hygiene and Tropical Medicine.^22^ Appendix 2 describes the implementation and validation of that algorithm for the present analysis. Duration of T2DM was defined by the difference between the index date and first recording of T2DM in the patient’s CPRD record from January 2000.

Patients’ IMD was divided into five groups defined by quintiles from least (5) to most (1) deprived. For CVD risk factors, including laboratory results and BMI, the most recent measurement within two years prior to the index date was used; if none were available in this window the value was set to missing. Presence of a current prescription of any medication was defined using the algorithm developed by Farmer *et al*.^23^ This estimates the days supply of a prescription using any data available on the quantity of pills and the daily dose; or number of days prescribed; or the average gap between previous prescriptions. If none of these are available a 28 day prescription length is assumed. If the most recent prescription date plus two times the estimated days supply falls after the index date, then the prescription is considered current. People with a current prescription for dapagliflozin, empagliflozin, canagliflozin, or ertugliflozin were counted as having a current SGLT2 inhibitor.

### Descriptive and statistical analysis

The demographic and clinical characteristics of the prevalent population with T2DM in September 2023 were summarised using counts, percentages, and means, stratified by comorbidity (CVD risk score category, and CVD and HF status).

Uptake of SGLT2 inhibitors in each CVD group was defined the percentage of the population with a current prescription on the index date. Uptake was calculated overall and stratified by patient characteristics.

The association between patient characteristics and odds of a current SGLT2 inhibitor prescription was then evaluated using logistic regression. First, one model was fitted to all people with either CVD or HF, with categorical variables for comorbidity status (CVD and CKD status) and all patient characteristics. Then, separate models were fitted for each CVD and CVD risk group (including people without CVD), each model including CKD status and all patient characteristics as covariables, to assess for heterogeneity in the association between patient characteristics and uptake between the different subpopulations. A 6-category T2DM duration variable was used to better control for any confounding from this. The analysis was conducted in STATA 16; all analysis code is available online (Appendix 1).

## RESULTS

### Baseline characteristics

#### Demographics

Initially 787,152 patients with prevalent T2DM and no record of any other kind of a diabetes, a research-eligible record, and one year history with their GP on 1^st^ September 2023 were extracted for analysis. Of these, 587,826 were over 18 years age and could be linked to hospital inpatient admissions and IMD category. These comprised the final study cohort included in the analysis (Table 1).

**Table 1.**
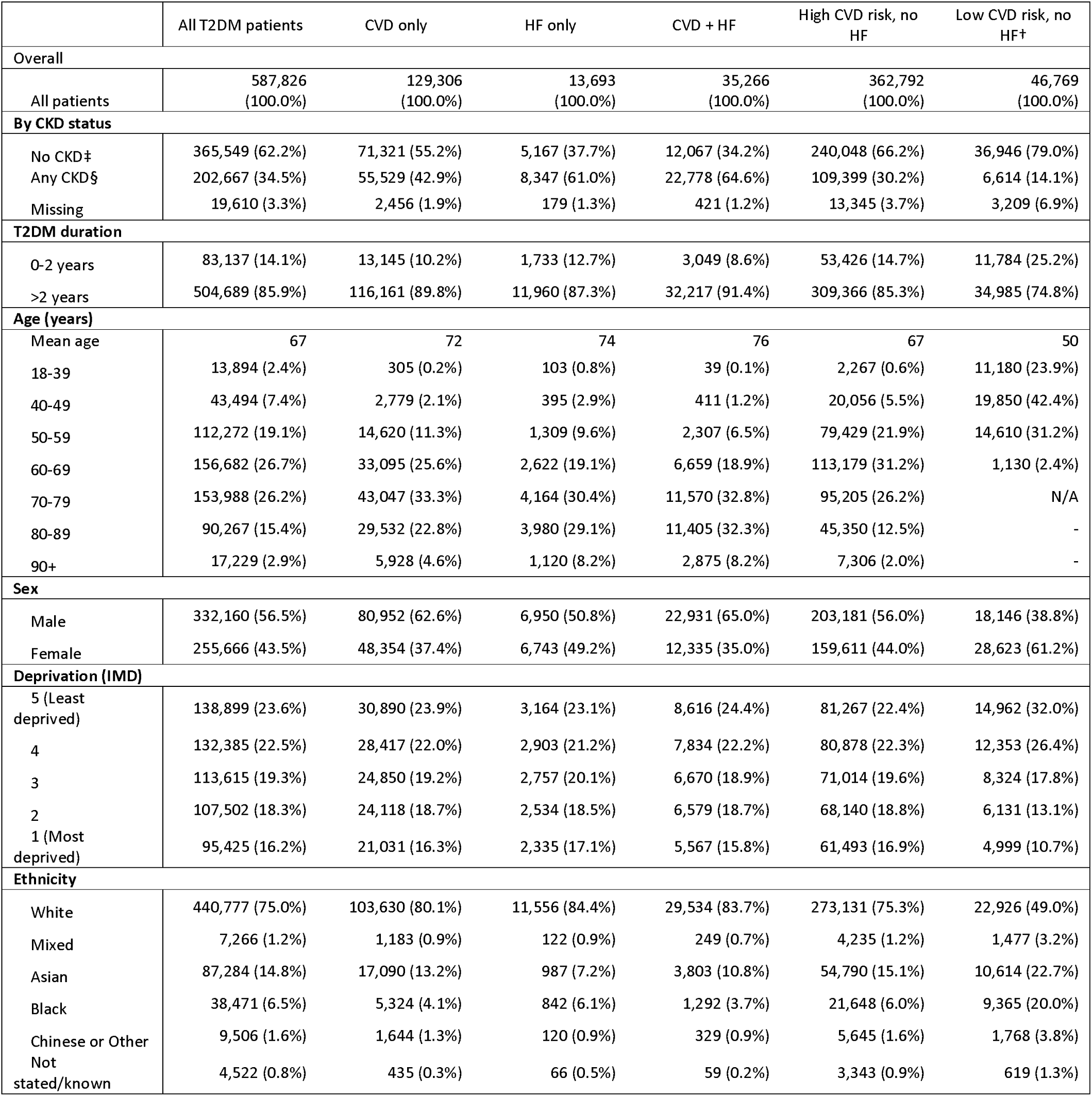
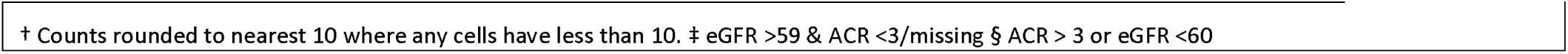
Clinical and demographic characteristics of people in CPRD with prevalent T2DM by cardiovascular disease comorbidity status, September 2023.

|  | All T2DM patients | CVD only | HF only | CVD + HF | High CVD risk, no HF | Low CVD risk, no HF† |
| --- | --- | --- | --- | --- | --- | --- |
| Overall |  |  |  |  |  |  |
| All patients | 587,826 (100.0%) | 129,306 (100.0%) | 13,693 (100.0%) | 35,266 (100.0%) | 362,792 (100.0%) | 46,769 (100.0%) |
| **By CKD status** |  |  |  |  |  |  |
| No CKD‡ | 365,549 (62.2%) | 71,321 (55.2%) | 5,167 (37.7%) | 12,067 (34.2%) | 240,048 (66.2%) | 36,946 (79.0%) |
| Any CKD§ | 202,667 (34.5%) | 55,529 (42.9%) | 8,347 (61.0%) | 22,778 (64.6%) | 109,399 (30.2%) | 6,614 (14.1%) |
| Missing | 19,610 (3.3%) | 2,456 (1.9%) | 179 (1.3%) | 421 (1.2%) | 13,345 (3.7%) | 3,209 (6.9%) |
| **T2DM duration** |  |  |  |  |  |  |
| 0-2 years | 83,137 (14.1%) | 13,145 (10.2%) | 1,733 (12.7%) | 3,049 (8.6%) | 53,426 (14.7%) | 11,784 (25.2%) |
| >2 years | 504,689 (85.9%) | 116,161 (89.8%) | 11,960 (87.3%) | 32,217 (91.4%) | 309,366 (85.3%) | 34,985 (74.8%) |
| **Age (years)** |  |  |  |  |  |  |
| Mean age | 67 | 72 | 74 | 76 | 67 | 50 |
| 18-39 | 13,894 (2.4%) | 305 (0.2%) | 103 (0.8%) | 39 (0.1%) | 2,267 (0.6%) | 11,180 (23.9%) |
| 40-49 | 43,494 (7.4%) | 2,779 (2.1%) | 395 (2.9%) | 411 (1.2%) | 20,056 (5.5%) | 19,850 (42.4%) |
| 50-59 | 112,272 (19.1%) | 14,620 (11.3%) | 1,309 (9.6%) | 2,307 (6.5%) | 79,429 (21.9%) | 14,610 (31.2%) |
| 60-69 | 156,682 (26.7%) | 33,095 (25.6%) | 2,622 (19.1%) | 6,659 (18.9%) | 113,179 (31.2%) | 1,130 (2.4%) |
| 70-79 | 153,988 (26.2%) | 43,047 (33.3%) | 4,164 (30.4%) | 11,570 (32.8%) | 95,205 (26.2%) | N/A |
| 80-89 | 90,267 (15.4%) | 29,532 (22.8%) | 3,980 (29.1%) | 11,405 (32.3%) | 45,350 (12.5%) | - |
| 90+ | 17,229 (2.9%) | 5,928 (4.6%) | 1,120 (8.2%) | 2,875 (8.2%) | 7,306 (2.0%) | - |
| **Sex** |  |  |  |  |  |  |
| Male | 332,160 (56.5%) | 80,952 (62.6%) | 6,950 (50.8%) | 22,931 (65.0%) | 203,181 (56.0%) | 18,146 (38.8%) |
| Female | 255,666 (43.5%) | 48,354 (37.4%) | 6,743 (49.2%) | 12,335 (35.0%) | 159,611 (44.0%) | 28,623 (61.2%) |
| **Deprivation (IMD)** |  |  |  |  |  |  |
| 5 (Least deprived) | 138,899 (23.6%) | 30,890 (23.9%) | 3,164 (23.1%) | 8,616 (24.4%) | 81,267 (22.4%) | 14,962 (32.0%) |
| 4 | 132,385 (22.5%) | 28,417 (22.0%) | 2,903 (21.2%) | 7,834 (22.2%) | 80,878 (22.3%) | 12,353 (26.4%) |
| 3 | 113,615 (19.3%) | 24,850 (19.2%) | 2,757 (20.1%) | 6,670 (18.9%) | 71,014 (19.6%) | 8,324 (17.8%) |
| 2 | 107,502 (18.3%) | 24,118 (18.7%) | 2,534 (18.5%) | 6,579 (18.7%) | 68,140 (18.8%) | 6,131 (13.1%) |
| 1 (Most deprived) | 95,425 (16.2%) | 21,031 (16.3%) | 2,335 (17.1%) | 5,567 (15.8%) | 61,493 (16.9%) | 4,999 (10.7%) |
| **Ethnicity** |  |  |  |  |  |  |
| White | 440,777 (75.0%) | 103,630 (80.1%) | 11,556 (84.4%) | 29,534 (83.7%) | 273,131 (75.3%) | 22,926 (49.0%) |
| Mixed | 7,266 (1.2%) | 1,183 (0.9%) | 122 (0.9%) | 249 (0.7%) | 4,235 (1.2%) | 1,477 (3.2%) |
| Asian | 87,284 (14.8%) | 17,090 (13.2%) | 987 (7.2%) | 3,803 (10.8%) | 54,790 (15.1%) | 10,614 (22.7%) |
| Black | 38,471 (6.5%) | 5,324 (4.1%) | 842 (6.1%) | 1,292 (3.7%) | 21,648 (6.0%) | 9,365 (20.0%) |
| Chinese or Other | 9,506 (1.6%) | 1,644 (1.3%) | 120 (0.9%) | 329 (0.9%) | 5,645 (1.6%) | 1,768 (3.8%) |
| Not stated/known | 4,522 (0.8%) | 435 (0.3%) | 66 (0.5%) | 59 (0.2%) | 3,343 (0.9%) | 619 (1.3%) |

† Counts rounded to nearest 10 where any cells have less than 10. ‡ eGFR >59 & ACR <3/missing § ACR > 3 or eGFR <60

Most people were aged over 60 years (71.2%), 56.5% were male, and 75.0% were white. People with T2DM were more likely to live in deprived areas than the general population: 138,899 (23.6%) were living in the most deprived quintile in England compared to 95,425 (16.2%) living in the least deprived.

#### Cardiovascular disease and heart failure status

Cardiovascular disease was common: 129,306 (22.0%) had CVD without HF; 13,693 (2.3%) had HF without CVD; 35,266 (6.0%) had CVD and HF; 362,792 (61.7%) had neither CVD or HF but estimated 10 year CVD risk > 10%; and 46,769 (8.0%) had neither but CVD risk <10%.

#### Chronic kidney disease

In total 202,667 (34.5%) of the cohort had a record of comorbid CKD. Of these, 31,835 (15.7%) had stage 1 disease, 50,800 (25.1%) had stage 2 disease, 104,882 (51.8%) had stage 3 disease, and 15,150 (7.5%) had stage 4 or 5 disease (Appendix 3). A record of eGFR within 2 years prior to the index date was missing for 3.3% of the cohort.

### Uptake of SGLT2 inhibitors

Overall uptake of SGLT-2 inhibitors for people with T2DM in General Practice was 20.7%, varying between 19.5% and 30.5% according to CVD group (Table 2). Uptake was higher in patients with heart failure (29.4% and 30.5% in patients with HF with and without CVD respectively) and lower in patients without heart failure (19.5%, 19.9%, and 20.2% in the patients without HF but with current CVD, high CVD risk, and low CVD risk, respectively). In the unadjusted results, uptake was markedly higher for people diagnosed with T2DM over 2 years prior to the analysis index date compared to those diagnosed more recently; higher in men than women; and higher in people aged under 70 years than those older than 70 years. Smaller differences in unadjusted uptake were present according to CKD status, deprivation, and ethnicity.

**Table 2.** Percentage of people with prevalent T2DM and a current prescription for an SGLT-2 inhibitor, by cardiovascular disease status, September 2023.

|  | All T2DM patients | CVD, no HF | HF, no CVD | HF and CVD | High CVD risk | Low CVD risk |
| --- | --- | --- | --- | --- | --- | --- |
| **Overall** |  |  |  |  |  |  |
| All patients | 20.7% | 19.5% | 29.4% | 30.5% | 19.9% | 20.2% |
| **CKD status** |  |  |  |  |  |  |
| No CKD | 20.9% | 20.7% | 29.8% | 31.6% | 20.4% | 20.1% |
| Any CKD | 21.8% | 18.5% | 29.7% | 30.3% | 20.7% | 27.8% |
| Missing | 6.5% | 7.6% | 8.4% | 9.0% | 6.4% | 5.4% |
| **T2DM duration** |  |  |  |  |  |  |
| 0-2 years | 8.3% | 7.8% | 27.9% | 25.5% | 6.8% | 8.6% |
| >2 years | 22.8% | 20.9% | 29.7% | 31.0% | 22.2% | 24.1% |
| **Age (years)** |  |  |  |  |  |  |
| 18-39 | 16.1% | 23.9% | 46.6% | 43.6% | 20.1% | 14.7% |
| 40-49 | 22.2% | 29.0% | 42.3% | 48.2% | 22.2% | 20.4% |
| 50-59 | 26.1% | 30.3% | 39.8% | 44.0% | 25.0% | 24.1% |
| 60-69 | 25.9% | 27.4% | 37.7% | 41.4% | 24.4% | 20.3% |
| 70-79 | 19.3% | 18.9% | 32.1% | 34.1% | 17.2% | N/A |
| 80-89 | 10.5% | 8.8% | 21.6% | 22.0% | 7.6% |  |
| 90+ | 4.4% | 2.4% | 9.8% | 11.0% | 2.6% |  |
| **Sex** |  |  |  |  |  |  |
| Male | 23.2% | 22.2% | 35.6% | 34.9% | 22.3% | 19.1% |
| Female | 17.5% | 15.1% | 23.1% | 22.4% | 17.0% | 20.9% |
| **Deprivation (IMD)** |  |  |  |  |  |  |
| 5 (Least deprived) | 20.9% | 20.4% | 28.7% | 30.7% | 20.0% | 19.3% |
| 4 | 20.7% | 19.7% | 30.1% | 30.5% | 20.0% | 19.5% |
| 3 | 20.5% | 19.3% | 28.5% | 29.5% | 19.8% | 20.3% |
| 2 | 20.9% | 19.1% | 30.8% | 31.3% | 20.1% | 22.4% |
| 1 (Most deprived) | 20.6% | 18.7% | 29.2% | 30.5% | 19.9% | 21.7% |
| **Ethnicity** |  |  |  |  |  |  |
| White | 21.0% | 18.9% | 29.5% | 30.0% | 20.3% | 22.7% |
| Mixed | 18.6% | 21.0% | 36.1% | 34.5% | 17.1% | 17.0% |
| Asian | 21.9% | 24.5% | 27.0% | 34.8% | 20.5% | 19.3% |
| Black | 16.5% | 16.5% | 29.8% | 26.9% | 15.5% | 16.3% |
| Chinese or Other | 18.6% | 20.7% | 35.8% | 35.3% | 17.0% | 17.6% |
| Not stated / known | 17.2% | 15.4% | 34.8% | 30.5% | 17.2% | 15.7% |

When stratified by age group, a consistent association between CVD group and SGLT2 inhibitor uptake was present, with uptake ordered from lowest to highest as follows: low CVD risk score, high CVD risk score, CVD only, HF only, CVD and HF (Appendix 5). In analysis stratified by CVD group and age, CKD was also consistently associated with higher uptake of SGLT2 inhibitors for patients in each age category with CVD or HF. In people aged 60-69 years with CVD, uptake was 26.2% in people without CKD and 31.5% in people with CKD; similar differences were present in other age groups.

### Modelling analysis of uptake of SGLT2 inhibitors

All clinical patient characteristics were significantly (p<0.01) associated with differences in the odds of having a current prescription for an SGLT2 inhibitor in a model for people with CVD or HF which mutually adjusted for all other patient characteristics, CVD group and CKD status (Table 3). CKD was associated with higher odds of uptake (OR: 1.22, 95% CI: (1.19, 1.25)), as was HF (OR: 2.15 (2.06, 2.24)) and combined CVD and HF (OR: 2.15 (2.09, 2.22)). Patients aged 70-79 years had considerably lower odds than patients aged 18-39 (OR: 0.41 (0.33, 0.51)), women had lower odds than men (OR: 0.67 (0.65, 0.68)), and people living in areas in the highest quintile of deprivation had lower odds than those in the least deprived (OR: 0.84 (0.81, 0.88)). Compared to people of White ethnicity, people of Asian ethnicity had higher odds of a current prescription (OR: 1.07 (1.04, 1.11)) whilst people Black ethnicity had lower odds (OR: 0.77 (0.72, 0.81)).

**Table 3.**
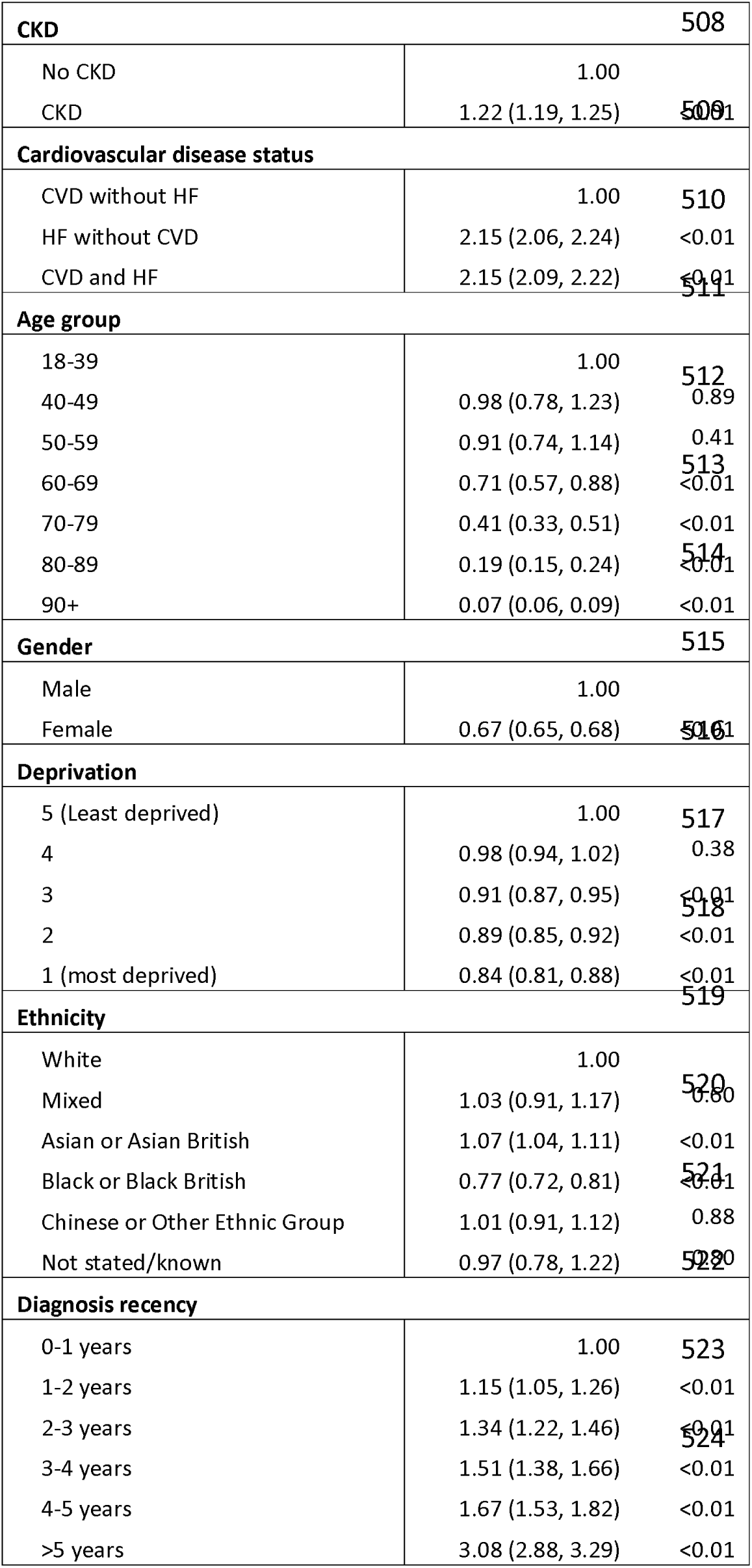
Logistic regression model for association between current SGLT2 inhibitor prescription and patient clinical and demographic characteristics, people with T2DM and either CVD or HF, September, 2023.

The same direction of association for each of the different patient characteristics in the overall population was also generally observed within each CVD and CVD risk group (Appendix 5, Figure 2). Higher age and female gender were consistently and strongly associated with lower odds in all groups, except the low CVD risk group, where odds were higher with higher age and only slightly lower for women (OR: 0.95 (0.90, 1.00)). Black ethnicity was associated with lower uptake in all groups, though the difference was greater in patients without HF. Asian ethnicity was associated with higher uptake in people with CVD. In patients with neither CVD nor HF, White ethnicity was associated with considerably higher uptake than any other ethnicity. CKD was associated with higher odds of a prescription in all subgroups (OR ranging between 1.17 and 1.47), while more recent diagnosis was consistently associated with lower odds except in patients without HF (Appendix Table 5).

**Figure 2.**
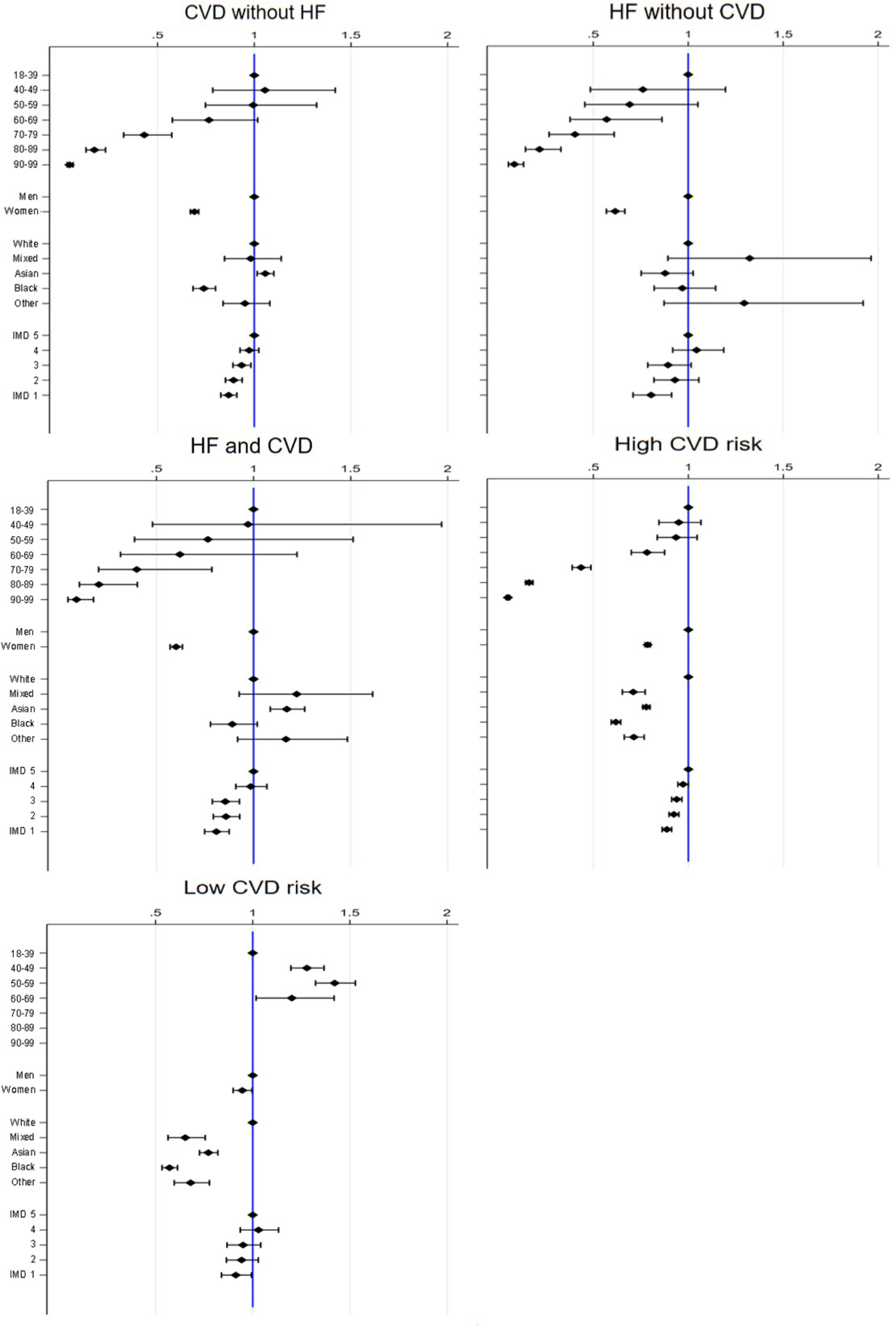
Odds ratios for association between patient characteristics and a current SGLT2 inhibitor prescription, by cardiovascular disease status, models adjusted for diagnosis recency and CKD stage, September 2023.

## Discussion

### Principal findings

The percentage of people with T2DM with a current SGLT2 inhibitor prescription remained relatively low by September 2023, 18 months after the 2022 NICE T2DM guideline update. Uptake was lower in people with comorbid CVD without HF (19.5%) and people at high CVD risk without HF (19.9%) than for people with HF (29.4%). In age-stratified results, uptake was higher with in people with comorbidities and greater CVD risk score: higher in people with CKD than those without CKD; higher in people with CVD than people at high CVD risk; and higher in people at high CVD risk than in those at low CVD risk.

In models which adjusted for patients’ characteristics and comorbidity status (CVD and CKD status), people aged 70-79 had less than half the odds of having a current prescription than people aged 18-39, and women had approximately two-thirds the odds of men. People with Black ethnicity consistently had lower odds of a current prescription than people with White ethnicity, ranging from 8-40% lower in the different groups, whilst people living in the most deprived areas had 9-10% lower odds than those living in the least deprived areas.

### Interpreting variation in uptake

#### Variation by comorbidity and NICE guidance type

Some of the variation in prescribing we report may be due to differences in the speed and extent of implementation of different NICE guidance, including Technology Appraisals on SGLT2 inhibitors for different indications between 2021 and 2023, and the 2022 T2DM guideline update (NICE NG28). Clinical considerations may partly explain some of the other inequalities we report.

NICE NG28 does not differentiate between CVD and HF in the strength of its recommendations, stating an SGLT2 inhibitor should be offered to people with T2DM ‘if they have chronic heart failure or established atherosclerotic cardiovascular disease’. However, we found prescribing was 50% higher in people with HF than in people with CVD without HF. This may be due to increases in prescribing up to September 2023 being driven by Technology Appraisals for SGLT2 inhibitors for HF (2021-2023), in addition to the NG28 (2022) guideline recommendations for their use for T2DM, while no Technology Appraisals or regulatory approval existed specifically for the use of SGLT2 inhibitors for CVD.^24^ Higher uptake for people with T2DM and HF may be due prescribing initiated outside general practice by HF specialists, and prescribing support from specialist HF and community teams.

Recommendations for SGLT2 inhibitors in groups not covered by a relevant Technology Appraisal for another condition appear to have been implemented to some extent in England. In age-stratified results, prescribing of SGLT2 inhibitors was higher in people with T2DM with elevated CVD risk scores than people with lower scores, and higher still for people with CVD. This ordering mirrors the strength of recommendations in NG28 in these different groups.

Differentiation by where the prescription was initiated or by primary reason for prescribing was not possible in the CPRD data. However, the higher rates of prescribing in non-HF cohorts with longer durations of diagnosis suggests that glycaemic management may have been a greater driver than cardiorenal protection in these groups, as additional glucose-lowering medicines are typically required over time. The increasing use of SGLT2 inhibitors in this period may also explain the concurrent downward trends in prescribing of sulphonylureas.^25^

#### Variation by age, gender, ethnicity, and deprivation

We report lower prescribing of SGLT2 inhibitors with higher age, particularly after age 70 years. This could be partly due to concern about adverse outcomes relating to SGLT2 inhibitor therapy, the avoidance of polypharmacy, and caution in the context of frailty. Differences by gender may be at least partially due to adverse effects including genital mycotic infections and recurrent UTIs being more common in women. However, it is notable that similar gender inequalities have also been reported for drugs where these are not a concern.^26^ The 2026 NICE T2DM guideline update cautions the use of SGLT2 inhibitors in people with T2DM and frailty, but highlights the sparsity of evidence for this population,^27^ while analysis of individual patient data indicates that frail patients are underrepresented in RCTs and may have poorer outcomes.^28^

The higher odds of a current prescription in Asian people may reflect awareness of higher cardiovascular risk in this population. Although lower prescribing in people with Black ethnicity could be partly due to concerns about DKA with these medicines,^29^ it is more likely that lower uptake in Black people, along with lower prescribing in areas of higher deprivation, relate to systemic barriers in accessing healthcare including differential provisional as well as financial, cultural and psychosocial factors. However, more granular analysis of patient records as well as qualitative research in primary care would be needed to draw any conclusions on this.

### Population health benefits of higher uptake of SGLT2 inhibitors

With the loss of exclusivity of the patent for dapagliflozin the UK, generic dapagliflozin has recently launched and there has been 82% price reduction in the NHS tariff between July 2025 and February 2026. The price reductions imply substantial population health benefits can now be gained from higher uptake whilst limiting displaced resources within the wider NHS.

### Comparison to other studies on SGLT2 inhibitor uptake in the UK

To our knowledge no previous studies have reported uptake of SGLT2 inhibitors for T2DM up to 2023. Volumes of GP prescriptions for dapagliflozin (for any indication) increased in England approximately 80% between September 2023 and August 2025.^30^ Applying this increase to overall uptake in the present study (21%), implies it could have reached approximately 37% in people with T2DM by August 2025.

Seghezzo *et al* reported large increases in uptake of SGLT2 inhibitors in people with HF with reduced ejection from approximately 8% in January 2022 to over 25% by May 2024 in UK General Practice, with no indication of a plateau by 2024, and higher prescribing in people with comorbid T2DM.^26^ Another large UK General Practice study on CKD up to December 2022 reported that 62.8% of people with CKD had guideline-directed indication for SGLT2 inhibitor, but uptake in that cohort with CKD and T2DM was only 22%.^31^ These results are consistent with those in our subgroups of T2DM patients with comorbid HF and CKD.

### Comparison to other studies on SGLT2 inhibitor uptake internationally

Gagnon *et al* evaluated uptake of SGLT2 inhibitors in patients with T2DM and comorbid HF, separately in a specialised heart function clinic and in all adults in Alberta, Canada, between 2018 and 2022.^32^ In the specialised clinic cohort it reached 49.1% by March 2022, whilst in the whole province it reached 26.4% by January 2022. In one population-based study of adults with T2DM hospitalised with HF in New South Wales in Australia, uptake of SGLT2 inhibitors had reached 13.2% by mid-2021, with lower uptake reported in older age groups, people with higher frailty, and, in contrast to our analysis, people with CKD.^33^ Another analysis of 114,000 adults with T2DM in primary care in Australia reported uptake of 13.6% in people without CKD and 14.4% in people with CKD in 2020-2021.^34^

### Limitations

Evaluation of the QRISK2 algorithm we applied indicated that up to 20% of people estimated as having a low CVD risk score (QRISK2 <10%) could have had a QRISK2 of greater than 10% (Appendix 2). This may lead to some under-estimation of the difference in uptake between people with high and low CVD risk. Similarly, some people in CPRD with CVD and HF may have been misclassified as not having them due to linked hospital data only being available to March 2021,^35^ which may result underestimation of uptake differences between groups. Similarly, CKD stage may be misclassified in some instances by exclusion of eGFR measurements more than two years prior to the analysis index date, though only 3% of patients were affected by this. Our definition of current prescription included a grade period after the end of the estimate days supply, which may result in slight overestimation of uptake. However, by nature of the data we could not explore dispensations of the prescriptions, and actual adherence to treatment will be lower than our uptake results.

We did not evaluate how many people had previously initiated but then stopped an SGLT2 inhibitor, would be unsuitable for treatment, or assess for polypharmacy in our population. These factors will explain a proportion of the patients not prescribed one. In the Canadian cohort study of T2DM and HF Gagnon *et al* reported that 62.4% of people met the eligibility criteria for trials of SGLT2 inhibitors,^32^ and in another UK cohort study Forbes *et al* reported that 62.8% of patients with T2DM and CKD were indicated for SGLT2 inhibitor therapy.^31^ Further research would be needed to identify a plausible ceiling for uptake for the people in our analysis.

## Conclusion

We found that by September 2023 prescribing of SGLT2 inhibitors remained relatively low for people with T2DM recommended them in recent NICE technology appraisals and guidelines in England. Important inequalities by gender, ethnicity, and deprivation were present. Comparing our results to data on national trends on SGLT2 inhibitor pharmacy dispensations up to 2025, there are encouraging signs that uptake has risen. However, these data suggest that a substantial opportunity to increase prescribing may still remain in 2026, though further surveillance of primary care prescribing for T2DM is needed to verify this.

A major update to the NICE T2DM guideline in February 2026 recommended that all T2DM patients, regardless of their CVD risk, should be offered both metformin and an SGLT2 inhibitor. This expands the eligible population to include all the clinical subgroups considered in the present analysis. Healthcare systems and policy makers should consider case-finding and quality improvement interventions, such as pay-for-performance mechanisms, to ensure comprehensive and equitable access in all these groups.

## Contribution to authorship

The study was conceived by LO, PM, and designed by all the authors. PM and JW did the data and statistical analysis with support from RW and EY; JW and MR produced the code lists. All authors assisted with the interpretation of results. PM wrote the manuscript with substantial input and revisions from all other authors. The corresponding author attests that all listed authors meet authorship criteria and that no other meeting the criteria have been omitted.

## Acknowledgements, funding, and disclaimer

PM, JW, MR, JH, RW, EY, SR, MW, LSR and LO are employees of the National Institute for Health and Care Excellence. AA is supported by funding from NIHR Oxford Biomedical Research Centre. The authors are solely responsible for any errors or omissions. The opinions expressed in this article are those of the authors and do not necessarily reflect the position of their affiliations.

## Disclosure of interests

DJC has received investigator-initiated grants from Astra Zeneca and Novo Nordisk, support for education from Perspectum and consultancy fees from Madrigal with any financial renumeration from pharmaceutical company consultation made to the University of Liverpool. All other authors declare no competing interests.

## Sharing of data and results

No additional data are available; the authors are not the data controllers and do not have permission to share the data.

## Transparency statement

PM had full access to all the data in the study, and affirms that the manuscript is an honest, accurate, and transparent account of the study being reported; that no important aspects of the study have been omitted; and that there are no discrepancies between the study conducted and the study as originally planned.

## Ethical approval

This study was based on data from the Clinical Practice Research Datalink obtained under licence from the UK Medicines and Healthcare products Regulatory Agency. The study was approved by the CPRD Research Data Governance process (ID: 23_003530).

## Notes

### Author Declarations

This study was based on data from the Clinical Practice Research Datalink obtained under licence from the UK Medicines and Healthcare products Regulatory Agency. The study was approved by the Clinical Practice Research Datalink Research Data Governance process (ID: 23_003530).

### Summary of Updates

Reformatted with some caveats about study limitations added, clarification of handling of missing ACR measurements, and change of title from "cohort" to "cross sectional" as the primary outcome of the analysis is cross sectional

