## Supplemental materials for "SGLT2 inhibitor use in type 2 diabetes in England: a population-based cross-sectional study of uptake of NICE guidance"

### APPENDICES

**Appendix 1.** Codelists used for identification of cohort and outcomes in the CPRD-HES dataset, and STATA code for statistical analysis.

All are stored at https://github.com/NICE-Data-and-Analytics in the repository "NICE_T2DM_project".

**Appendix 2.** Identification of type 2 diabetes people at low and high risk of cardiovascular disease (CVD) using the CPRD General Practice database.

The QRISK2 algorithm is used in General Practice in the UK to estimate patient's 10-year CVD risk as part of CVD primary prevention,[23] with a score over 10% indicating high risk. Analysis of CPRD indicated that a timely (within 2 months either way) QRISK2 was only recorded for 28% of newly diagnosed T2DM people in 2023, with a smaller percentage expected for the prevalent population on 1st September 2023.

It was determined that the best approach for cohort selection was to directly estimate QRISK2 at 1st September 2023 for all people without a timely QRISK2 record, using relevant clinical records in patient's GP records. Herrett *et al* previously created a STATA programme to estimate QRISK2 using CPRD Aurum records,[24] and published the code on GitHub.[42] In brief, the programme searches for relevant diagnoses and most recent clinical measurements in CPRD, and estimates QRISK2 using these. Where an item needed to calculate QRISK2 for a patient is missing, population-average results are substituted. We applied this programme to our CPRD Aurum extracts. CPRD drug issue and observation records prior to September 2023 were used for the calculation, but patient's Townsend scores were not available.

We evaluated the concordance of programme-estimated QRISK2 on 1st September 2023 and actual recorded scores for 387 people who also had GP-recorded QRISK2 on 1st September 2023 (Appendix 2 Figure 1). Concordance in risk category (low (<10%) or high (>10%)) between programme-estimated and GP-recorded QRISK2 scores was 92.0% (Appendix 2 Table 1). However, people with GP-recorded low risk were more likely to be categorised differently by the programme: the programme classified only 77.0% of these people as also being low risk, whereas it classified 95.5% of people with GP-recorded high risk as high risk.

The QRISK2 programme gave a higher mean QRISK2 than GP entered QRISK2: 24.0% compared to 22.4%. These results mirror the report from Herrett *et al* on an evaluation in CPRD GOLD: they reported the average programme QRISK2 was 0.4% higher than the GP-recorded, but notably, it was 1% higher for people with type 2 diabetes and 4.5% higher for people with CKD.[42] Within our sample, in those instances where the GP recorded QRISK2 was <10% whilst the programme estimated was >10%, the median programme-estimated QRISK2 was 12.9% and the mean was 14.3%, indicating that whilst these people were above the high risk threshold, they were typically also close to it.

Appendix 2 Figure 1. Concordance of GP-recorded and programme estimated QRISK2 scores on 1st September 2023 (line at x=y).

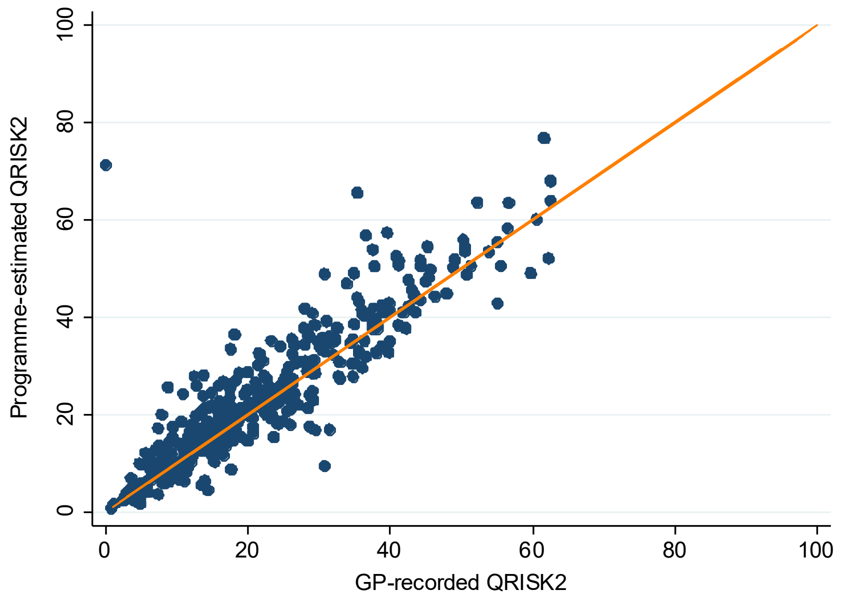


Appendix 2 Table 1. Concordance of CVD risk categorisation from GP-recorded and programme-estimated QRISK2 scores on 1st September 2023.

| | GP-recorded < 10% | GP recorded > 10% | Total |
|---|---|---|---|
| **Programme estimated < 10%** | 57 (80.3%) | 14 (19.7%) | 71 (100.0%) |
| **Programme estimated > 10%** | 17 (5.3%) | 299 (94.6%) | 316 (100.0%) |
| **Overall misclassification: 31/387 (8.0%)** | | | |

Additional initial analysis was done to evaluate the concordance of SGLT2 inhibitor uptake statistics between patients with algorithm-estimated high and low CVD risk (the whole analysis cohort of 589,000 patients) and patients with GP-recorded high and low CVD risk (including the 59,000 patients who had a GP-recorded QRISK2 score between January and September 2023). The results show that, within each age group, uptake was generally similar between people with GP-recorded high risk and algorithm-estimated high risk (Appendix 2 Table 2), whilst for patients with low CVD risk aged 40-70 years uptake was 2-5% higher for people with algorithm-estimated low risk compared to GP recorded low risk. The latter finding may be due to more recent T2DM diagnosis in people with a GP-recorded QRISK2 in 2023, as assessment is recommended following diagnosis, and this could potentially explain the lower uptake. Further consideration of the association between more recent diagnosis and lower uptake is presented the discussion.

Appendix 2 Table 2. Uptake of SGLT2 inhibitors by age group and CVD risk category (<10%, >10%, either estimated algorithmically or taken from GP recording).

| **Age (years)** | High CVD risk: algorithm estimated, whole cohort | High CVD risk: GP recorded between Jan '23 and Sep '23 | Low CVD risk: algorithm estimated, whole cohort | Low CVD risk: GP recorded between Jan '23 and Sep '23 |
|---|---|---|---|---|
| 18-39 | 20.1% | 21.0% | 14.7% | 18.0% |
| 40-49 | 22.2% | 20.7% | 20.4% | 18.4% |
| 50-59 | 25.0% | 22.5% | 24.1% | 19.0% |
| 60-69 | 24.4% | 22.4% | 20.3% | 15.0% |
| 70-79 | 17.2% | 19.4% | | |
| 80-89 | 7.6% | 12.0% | | |

**Appendix 3.** Distribution of eGFR and ACR measurements in the population with prevalent T2DM in CPRD Aurum, September 2023.

| **eGFR** | ACR <3 | ACR 3-30 | ACR > 30 | ACR Missing | Total |
|---|---|---|---|---|---|
| >90 | 95,045 | 29,121 | 2,714 | 60,283 | 187,163 (31.8%) |
| 60-89 | 137,528 | 46,200 | 4,600 | 72,693 | 261,021 (44.4%) |
| 45-59 | 31,080 | 17,221 | 2,808 | 17,141 | 68,250 (11.6%) |
| 30-44 | 12,851 | 11,639 | 2,961 | 9,181 | 36,632 (6.2%) |
| 15-29 | 2,327 | 4,331 | 2,083 | 3,593 | 12,334 (2.1%) |
| <15 | 60 | 375 | 570 | 1,811 | 2,816 (0.5%) |
| Missing | 695 | 238 | 39 | 18,638 | 19,610 (3.3%) |
| Total | 279,586 | 109,125 | 15,775 | 183,340 | 587,826 (100.0%) |

**Appendix 4.** Percentage of people with a current prescription for an SGLT-2 inhibitor, in patients with T2DM and heart failure or CVD, by CKD status, September 2023.

| | CVD only | | HF only | | CVD + HF | |
|---|---|---|---|---|---|---|
| | CKD | No CKD | CKD | No CKD | CKD | No CKD |
| **Overall** | | | | | | |
| All patients | 18.5% | 20.7% | 29.7% | 29.8% | 30.3% | 31.6% |
| **T2DM duration** | | | | | | |
| 0-2 years | 28.7% | 47.5% | 52.5% | 52.6% | 48.3% | 48.0% |
| >2 years | 19.3% | 22.7% | 29.8% | 30.3% | 30.6% | 32.7% |
| **Age (years)** | | | | | | |
| 18-39 | 31.0% | 23.1% | 69.2% | 39.7% | 18.2% | 60.0% |
| 40-49 | 36.4% | 27.9% | 44.5% | 41.3% | 51.6% | 49.6% |
| 50-59 | 36.1% | 29.5% | 46.4% | 37.6% | 44.8% | 44.8% |
| 60-69 | 31.5% | 26.2% | 38.6% | 37.9% | 43.7% | 40.4% |
| 70-79 | 20.7% | 17.7% | 35.3% | 27.5% | 36.9% | 29.6% |
| 80-89 | 9.6% | 7.8% | 23.8% | 14.9% | 23.8% | 16.3% |
| 90+ | 2.6% | 2.2% | 10.8% | 5.3% | 11.6% | 8.2% |
| **Sex** | | | | | | |
| Male | 21.8% | 22.9% | 36.2% | 35.5% | 35.1% | 35.3% |
| Female | 13.6% | 16.8% | 23.7% | 22.7% | 22.6% | 22.4% |
| **Deprivation (IMD)** | | | | | | |
| 5 (Least deprived) | 19.4% | 21.6% | 28.9% | 29.0% | 29.8% | 33.1% |
| 4 | 19.0% | 20.8% | 29.8% | 31.4% | 30.1% | 31.9% |
| 3 | 18.7% | 20.2% | 29.6% | 27.3% | 29.8% | 29.6% |
| 2 | 18.0% | 20.4% | 30.3% | 32.3% | 31.3% | 31.7% |
| 1 (Most deprived) | 17.2% | 20.2% | 29.8% | 28.8% | 30.8% | 30.6% |
| **Ethnicity** | | | | | | |
| White | 17.5% | 20.3% | 29.5% | 30.0% | 29.9% | 30.9% |
| Mixed | 22.2% | 21.5% | 31.5% | 44.7% | 34.4% | 35.3% |
| Asian | 25.7% | 24.2% | 28.0% | 26.4% | 34.2% | 37.0% |
| Black | 16.9% | 16.8% | 32.7% | 25.8% | 27.5% | 27.8% |
| Chinese or Other | 21.1% | 21.5% | 33.3% | 42.0% | 33.5% | 39.7% |
| Not stated / known | 20.6% | 14.2% | 34.2% | 40.0% | 31.3% | 33.3% |

**Appendix 5** Results from different logistic regression models for the association between current SGLT2 inhibitor prescription and patient clinical and demographic characteristics, people with T2DM and different comorbidity status, Sep 2023.

| | CVD, no HF | | HF, no CVD | | HF and CVD | | High CVD risk | | Low CVD risk | |
|---|---|---|---|---|---|---|---|---|---|---|
| | Odds ratio (95% CI) | P-value | Odds ratio (95% CI) | P-value | Odds ratio (95% CI) | P-value | Odds ratio (95% CI) | P-value | Odds ratio (95% CI) | P-value |
| **CKD** | | | | | | | | | | |
| No CKD | 1.00 | | 1.00 | | 1.00 | | 1.00 | | 1.00 | |
| CKD | 1.17 (1.14, 1.21) | <0.01 | 1.37 (1.26, 1.49) | <0.01 | 1.32 (1.25, 1.39) | <0.01 | 1.27 (1.24, 1.29) | <0.01 | 1.47 (1.38, 1.56) | 0.00 |
| **Age group** | | | | | | | | | | |
| 18-39 | 1.00 | | 1.00 | | 1.00 | | 1.00 | | 1.00 | |
| 40-49 | 1.07 (0.80, 1.43) | 0.66 | 0.77 (0.49, 1.21) | 0.25 | 1.05 (0.53, 2.11) | 0.88 | 0.94 (0.84, 1.05) | 0.28 | 1.27 (1.19, 1.36) | <0.01 |
| 50-59 | 0.99 (0.75, 1.32) | 0.96 | 0.68 (0.45, 1.04) | 0.07 | 0.83 (0.42, 1.62) | 0.58 | 0.92 (0.82, 1.03) | 0.13 | 1.41 (1.31, 1.51) | <0.01 |
| 60-69 | 0.75 (0.57, 1.00) | 0.05 | 0.57 (0.38, 0.86) | 0.01 | 0.69 (0.35, 1.34) | 0.28 | 0.75 (0.67, 0.84) | <0.01 | 1.17 (0.99, 1.38) | 0.07 |
| 70-79 | 0.41 (0.31, 0.55) | <0.01 | 0.42 (0.28, 0.63) | <0.01 | 0.46 (0.24, 0.90) | 0.02 | 0.40 (0.36, 0.45) | <0.01 | 1.00 (0.00, 0.00) | <0.01 |
| 80-89 | 0.16 (0.12, 0.22) | <0.01 | 0.23 (0.15, 0.35) | <0.01 | 0.25 (0.13, 0.49) | <0.01 | 0.14 (0.13, 0.16) | <0.01 | 1.00 (0.00, 0.00) | <0.01 |
| 90+ | 0.04 (0.03, 0.06) | <0.01 | 0.09 (0.06, 0.15) | <0.01 | 0.11 (0.06, 0.22) | <0.01 | 0.04 (0.04, 0.05) | <0.01 | 0.95 (0.90, 1.00) | 0.04 |
| **Gender** | | | | | | | | | | |
| Male | 1.00 | | 1.00 | | 1.00 | | 1.00 | | 1.00 | |
| Female | 0.69 (0.67, 0.71) | <0.01 | 0.62 (0.58, 0.67) | <0.01 | 0.61 (0.58, 0.64) | <0.01 | 0.78 (0.77, 0.79) | <0.01 | 0.95 (0.90, 1.00) | 0.04 |
| **Deprivation** | | | | | | | | | | |
| 5 (Least deprived) | 1.00 | | 1.00 | | 1.00 | | 1.00 | | 1.00 | |
| 4 | 0.97 (0.93, 1.02) | 0.27 | 1.03 (0.91, 1.17) | 0.61 | 0.99 (0.91, 1.07) | 0.73 | 0.97 (0.94, 1.00) | 0.05 | 1.03 (0.94, 1.13) | 0.53 |
| 3 | 0.93 (0.89, 0.98) | 0.01 | 0.89 (0.79, 1.01) | 0.08 | 0.86 (0.79, 0.93) | <0.01 | 0.94 (0.91, 0.97) | <0.01 | 0.95 (0.87, 1.04) | 0.29 |
| 2 | 0.89 (0.85, 0.94) | <0.01 | 0.93 (0.82, 1.05) | 0.24 | 0.86 (0.79, 0.93) | <0.01 | 0.92 (0.90, 0.95) | <0.01 | 0.94 (0.87, 1.03) | 0.19 |
| 1 (most deprived) | 0.87 (0.83, 0.91) | <0.01 | 0.80 (0.70, 0.90) | <0.01 | 0.81 (0.75, 0.87) | <0.01 | 0.89 (0.86, 0.91) | <0.01 | 0.91 (0.84, 1.00) | 0.04 |
| **Ethnicity** | | | | | | | | | | |
| White | 1.00 | | 1.00 | | 1.00 | | 1.00 | | 1.00 | |
| Mixed | 0.97 (0.84, 1.13) | 0.70 | 1.22 (0.83, 1.80) | 0.31 | 1.15 (0.87, 1.51) | 0.32 | 0.71 (0.65, 0.77) | <0.01 | 0.65 (0.56, 0.75) | <0.01 |
| Asian or Asian British | 1.06 (1.02, 1.10) | 0.00 | 0.83 (0.72, 0.97) | 0.02 | 1.12 (1.04, 1.21) | <0.01 | 0.79 (0.77, 0.81) | <0.01 | 0.78 (0.73, 0.83) | <0.01 |
| Black or Black British | 0.72 (0.67, 0.78) | <0.01 | 0.92 (0.78, 1.09) | 0.33 | 0.82 (0.72, 0.94) | <0.01 | 0.61 (0.58, 0.63) | <0.01 | 0.57 (0.53, 0.61) | <0.01 |
| Chinese or Other Ethnic Group | 0.96 (0.84, 1.08) | 0.48 | 1.24 (0.84, 1.82) | 0.29 | 1.12 (0.89, 1.43) | 0.33 | 0.72 (0.67, 0.78) | <0.01 | 0.69 (0.60, 0.78) | <0.01 |
| Not stated/known | 0.90 (0.68, 1.19) | 0.47 | 1.29 (0.76, 2.21) | 0.34 | 0.96 (0.54, 1.71) | 0.89 | 0.89 (0.81, 0.98) | 0.02 | 0.87 (0.69, 1.10) | 0.25 |
| **Diagnosis recency** | | | | | | | | | | |
| 0-1 years | 1.00 | | 1.00 | | 1.00 | | 1.00 | | 1.00 | |
| 1-2 years | 1.37 (1.21, 1.57) | <0.01 | 0.98 (0.79, 1.21) | 0.84 | 0.93 (0.78, 1.10) | 0.37 | 1.79 (1.67, 1.91) | <0.01 | 1.64 (1.44, 1.87) | <0.01 |
| 2-3 years | 1.74 (1.54, 1.98) | <0.01 | 0.83 (0.66, 1.04) | 0.10 | 1.11 (0.94, 1.30) | 0.23 | 2.48 (2.31, 2.65) | <0.01 | 2.19 (1.92, 2.49) | <0.01 |
| 3-4 years | 2.13 (1.88, 2.43) | <0.01 | 0.87 (0.68, 1.11) | 0.27 | 1.08 (0.91, 1.28) | 0.37 | 2.99 (2.79, 3.21) | <0.01 | 2.89 (2.52, 3.30) | <0.01 |
| 4-5 years | 2.63 (2.33, 2.97) | <0.01 | 1.10 (0.88, 1.38) | 0.41 | 0.87 (0.74, 1.03) | 0.12 | 3.63 (3.40, 3.87) | <0.01 | 3.23 (2.84, 3.68) | <0.01 |
| >5 years | 5.44 (4.93, 6.01) | <0.01 | 1.29 (1.10, 1.51) | <0.01 | 1.42 (1.26, 1.61) | <0.01 | 8.03 (7.61, 8.48) | <0.01 | 5.90 (5.32, 6.56) | <0.01 |
